# COVID-19 Trends in Florida – August 10 – November 14, 2020

**DOI:** 10.1101/2020.11.24.20235861

**Authors:** Rebekah D. Jones, Jacob E. Romer

**Affiliations:** The Covid Monitor, Florida; Florida COVID Action, Florida

## Abstract

Limited research on the prevalence and characteristics of SARS-CoV-2 (COVID-19) in K-12 environments has led to a flurry of speculative, qualitative, non-data-driven analysis with potentially dangerous implications for public health policy. Twenty-four US states provide, at a minimum, outbreak data in K-12 schools. Student enrollment across Florida’s 67 districts totals more than 2.67 million, with five of the ten most populated districts in the country in the state. This study uses Florida as a case study in COVID-19 trends in schools. With no state-wide mask mandate, varying levels of virtual-instruction participation, and a range of rural, suburban, and urban environments, Florida represents the wide range of learning environments and COVID-19 approaches to mitigation observed across the country. The Covid Monitor began tracking case data in K-12 schools in July 2020, and has since reported more than 200,000 confirmed cases of the virus in the K-12 environment nationwide – the largest date repository for cases in schools. The Covid Monitor’s national database allows for analysis of the characteristics and trends of cases in schools over time. The results may inform decisions about continued in-person and virtual learning access and promotion across the United States, as well as mitigation measures related to reopening policies across districts responsive to model results. These results also provide a baseline for monitoring trends and evaluating mitigation strategies.

**Table of Contents Summary:** Using the most comprehensive database of K-12 COVID-19 case data in the country, Florida provides clues for understanding student and staff cases in schools.

**What’s known on this subject:** Florida schools began reopening to in-person instruction in August and reported 15,393 student and staff cases of COVID-19 as of November 14, 2020. Incidence of COVID-19 cases in K-12 students and staff is of urgent public health concern.

**What this study adds:** COVID-19 cases reported in Florida schools were most influenced by community case rates and percent of students attending face-to-face. Student case rates were highest in high schools (4.5 per 1,000); staff case rates were highest in elementary levels (4.5-4.8).

## Background

Student enrollment across Florida’s 67 districts totals more than 2.67 million, with five of the ten most populated districts in the country in the state. Analysis of 3,451 laboratory-confirmed cases of SARS-CoV-2 in Florida K-12 schools during August 10 – October 3, 2020 might inform decisions about continued in-person and virtual learning access and promotion across the United States, as well as mitigation measures related to reopening policies across districts responsive to model results. These results also provide a baseline for monitoring trends and evaluating mitigation strategies.

Confirmed COVID-19 cases in K-12 settings were identified from district-level reporting in 41 of Florida’s 67 school districts, which combined represent 89% (2.4 million) of all K-12 students in the state. Thirty-five school districts provided COVID-19 case data as well as detailed data related to student and staff cases in varying grade levels, and enrollment totals for their district, including identification of students enrolled in face-to-face instruction. Together the selected counties with available data represented 90% (1.9 million) of all students in districts that were open to in-person instruction during the study period*. Approximately 55% of all students in the counties examined returned to in-person instruction during the report period, with 45% enrolled in virtual-learning programs or withdrawing from the district. The selected counties recorded 15,395 COVID-19 cases during the study period (Table 1).

**TABLE 1:** Confirmed cases of COVID-19 reported by Florida school districts, August 10 – October 1, 2020. ^†§¶^

| Grade Levels | Case Type |  |  | Total |
| --- | --- | --- | --- | --- |
|  | Student | Staff | *Not Specified* |  |
| Elementary | 473 | 351 | 130 | 954 |
| Middle | 221 | 141 | 36 | 398 |
| High | 591 | 158 | 134 | 883 |
| Other | 60 | 30 | 5 | 95 |
| *Not Specified* | 709 | 402 | 100 | 1211 |
| **Total** | **2054** | **1082** | **405** | **3541** |

<sup>†</sup>“Other” cases refer to alternative school programs, such as juvenile detention facilities, special education programs, or schools where enrollment by grade was not available. <sup>§</sup>“Not specified” indicates cases reported by a school district in which the role of student or staff was not defined. <sup>¶</sup>Grade levels were defined as elementary (PreK—grade 5), middle (grades 6—8), and high (grades 9—12).

Analysis of 15,395 laboratory-confirmed cases of SARS-CoV-2 (COVID-19) in Florida K-12 schools during August 10 – November 14, 2020 showed disproportional infection rates in students (71%) compared to teachers and staff (29%), consistent with data provided by other states. Staff cases represented the greatest share of cases in younger age-groups (40% in elementary) compared to older age groups (18% in high school).

A generalized linear model with Poisson error distribution and log link was used to model the total count of cases within school districts under 44 different conditions. The outcome of interest was the total counts of COVID-19 cases within a school district (in-person students and staff combined). Analyses were performed on two data sets: one from self-reporting of cases by school districts and one from the Florida Department of Health (Model A and Model B, respectively).

For Model A, three independent variables were used to model the count of total cases within a district: 1) percent in-person enrollment; 2) cases per 1,000 per week within the county population for the same time period; and 3) total district enrollment and staff (squared-root transformed). The estimated number of total cases and case rates over a 30-day period for a district with 25,000 students (Figure 1) considered community incidence rates ranging from 0.5 to 3.0 cases per 1,000 per week. To adjust for the varying exposure time, an offset term of log (days) was included in the model (see estimated regression coefficients in Table 2).

**TABLE 2:** Poisson regression coefficient estimates modelling counts of COVID-19 cases by district. ** ^††^

| MODEL A: Data Set 1 – from school district reports, varying time periods (n=32) |  |  |  |
| --- | --- | --- | --- |
| Coefficient | Estimate | Std. Error | z-value |
| Intercept | -2.331 | 0.180 | -12.9 |
| Percent in-person | 2.030 | 0.183 | 11.1 |
| Community Rate (1,000 per week) | 0.343 | 0.035 | 9.7 |
| Sqrt (District Enrollment + Staff) | 0.007 | 0.0002 | 34.2 |
| MODEL B. Data Set 2 – from FL DOH school report, 9/6/2020-10/3/2020 (n=49) |  |  |  |
| Coefficient | Estimate | Std. Error | z-value |
| Intercept | -6.497 | 0.440 | -14.8 |
| Percent in-person | 1.198 | 0.185 | 6.5 |
| Community Rate (1,000 per week) | 0.448 | 0.038 | 11.8 |
| Log (District Enrollment + Staff) | 0.881 | 0.028 | 30.9 |
| Days Open | 0.013 | 0.004 | 3.5 |

*\*\*Community case incidence was calculated based on laboratory—confirmed COVID-19 case data from the Florida Department of Health, retrieved October 10, 2020.*
*http://www11.doh.state.fl.us/comm/partners/covid19_report_archive/ †† County population data retrieved from the United States Census Bureau, American Community Survey, Population Statistics, December 2019. https://data.census.gov/cedsci/all?q=population%20florida%20counties*

**FIGURE 1:**
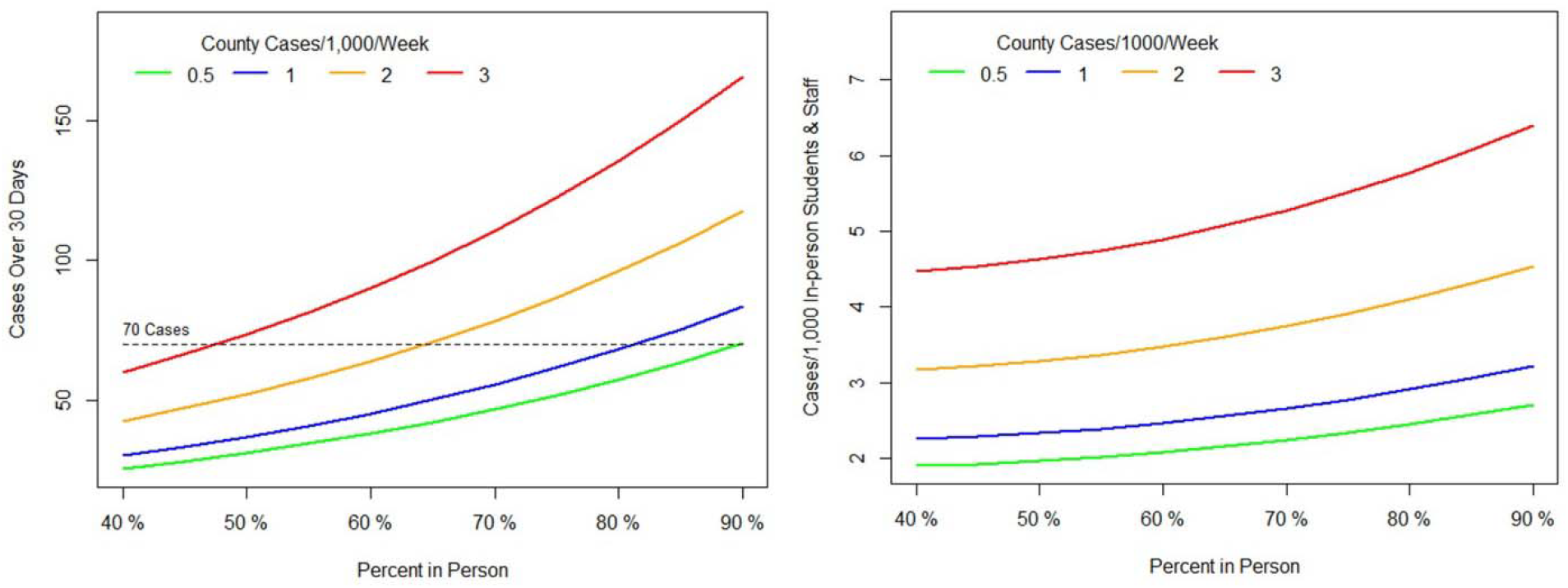
Influence of percent in-person attendance and COVID-19 community case rate on case totals and case incidence in K-12 school.

Cases related to schools reported by the Florida Department of Health (DOH) were used to model the effect of the same three variables on school incidence rate. Unlike the district-provided data, the DOH data was collected over a period of 28 days, from 9/6/2020 – 10/3/2020. The DOH report, which included data for all 67 Florida counties, under-reported the case data provided by 41 of the state’s districts by more than 1,238 confirmed cases. The DOH data used here for comparison purposes is not representative of case totals in K-12 schools in the state during the report period, but it is the only data made available by the state.

Only districts with known in-person enrollment were included in the state-portion of the analysis, and schools that had not yet begun in-person instruction were excluded (Miami-Dade and Broward). Each school district included in the analysis was open the entire period but for varying lengths of time prior to 9/6/2020. To adjust for this, ‘Days Open’ is included in the model and is calculated as the number of days between 9/6/2020 and the first day of school. The variable for total persons (district enrollment and staff) was log-transformed as opposed to the square-root in the previous model. R (version 4.0.2) was used to conduct all analyses.

## Results

During August 10 – October 1, 2020, average incidence (cases per 1,000 students enrolled in face-to-face instruction) in Florida high school students (4.5) was nearly twice that of elementary students (2.4). Overall staff rates (5.1) were more than twice the overall student rate (2.3). Staff case rates were highest at the elementary (4.5) and middle-school (4.8) level, and only lower than student incidence at the high-school level (4.2). The proportion of student to staff cases in Florida schools was closest in the elementary setting (59% students – 41% staff) compared to the high school setting (80% students – 20% staff).

A 1 percentage-point increase of in-person students (e.g. 60% to 61%) would have an estimated increase in district-wide student and staff case rates of about 2.1% (Model A, Table 2). For each increase in 1 case per 1,000 per week in the community, the average rate within schools is estimated to increase by more than 41%. For each increase of one unit of the squared-root of total persons (staff and in-person students), the expected rate increases by 0.7%.

The total number of expected cases in K-12 schools was lower for 90% in-person enrollment at low levels of community infection (0.5 cases per 1,000 per week) than for half that amount (48%) in-person enrollment at much higher community incidence rates (3.0 cases per 1,000 per week). Thus, the level of infection within the community appears to be the primary factor that influences rate of infections in school, with the percent of in-person students by enrollment total a significant secondary influencer on K-12 case incidence.

The results of the regression (Model B, Table 2) confirm the significant effect of both in-person percentage of students and community case rate on case incidence in schools. Differences in the coefficient estimates are likely due to different observation periods, the counties included in each analysis, and actual differences in the number of confirmed cases by DOH and the districts themselves. Despite the different model estimates, the conclusion is that lower in-person enrollment and community infection are important factors in the number of school cases.

## Discussion

Case incidence varies significantly between school grade levels and between students and staff. Staff rates are higher than student rates in all school environments except high schools. The rate of cases within schools is highly correlated with cases within a community, more than the size of the district by total enrollment. Percent enrollment in face-to-face instruction is a secondary influencer of case incidence rates in schools. For example, a 10 percentage-point increase in in-person enrollment would increase the school district’s case rate by 22.5%.

These findings can provide a baseline for monitoring national trends. A progressive, stratified reintroduction to in-person learning in communities with low case incidence rates could provide for a significantly lower risk of transmission in students, staff, and communities. Along with the implementation of mitigation methods proposed by the CDC, distance learning until community transmission is largely under control could have a profound impact on COVID-19 in the K-12 environment. Once community transmission reaches advised levels, a phased re-entry to in-person learning, especially at the high school level, could prevent further spread within the school setting.

## Data Availability

Data is available upon request, and at The Covid Monitor.com

https://github.com/TheCovidMonitor/Data

## Acknowledgements

The Covid Monitor provided data related to cases in K-12 school districts in Florida.

## References

1. Leeb RT, Price S, Sliwa S, et al. COVID-19 Trends Among School-Aged Children — United States, March 1–September 19, 2020. MMWR Morb Mortal Wkly Rep 2020;69:1410–1415. DOI: http://dx.doi.org/10.15585/mmwr.mm6939e2

2. United States Centers for Disease Control. 2020. Indicators for Dynamic School Decision-Making. https://www.cdc.gov/coronavirus/2019-ncov/community/schools-childcare/indicators.html

